# Belgian COVID-19 Mortality, Excess Deaths, Number of Deaths per Million, and Infection Fatality Rates (9 March — 28 June 2020)

**DOI:** 10.1101/2020.06.20.20136234

**Authors:** Geert Molenberghs, Christel Faes, Johan Verbeeck, Patrick Deboosere, Steven Abrams, Lander Willem, Jan Aerts, Heidi Theeten, Brecht Devleesschauwer, Natalia Bustos Sierra, Françoise Renard, Sereina Herzog, Patrick Lusyne, Johan Van der Heyden, Herman Van Oyen, Pierre Van Damme, Niel Hens

## Abstract

**Background:** COVID-19 mortality and its relation to excess deaths, the number of Deaths Per Million (DPM), Infection Fatality Rates (IFRs) and Case Fatality Rates (CFRs) are constantly being reported and compared for a large number of countries globally. These measures may appear objective, however they should be interpreted with the necessary care.

**Objective:** Scrutiny of COVID-19 mortality in Belgium over the period 9 March – 28 June 2020 (Weeks 11–26), using the relation between COVID-19 mortality and excess death rates, the number of deaths per million, and infection fatality rates.

**Methods:** The relation between COVID-19 reported mortality and excess death rates is evaluated by comparing publicly available COVID-19 mortality (2020) and the difference of observed and predicted overall mortality. Predictions are based on weekly averages of historical overall mortality data in Belgium (2009–2019). Deaths per million are evaluated using demographic data of the Belgian population (2020). The infection fatality rate is estimated using a delay distribution between infection and death. The number of infections in Belgium is estimated by a stochastic compartmental model, which uses hospitalisation data, serial serological survey data, and COVID-19 mortality data (2020) for calibration.

**Results:** In Belgium, 9621 COVID-19 related deaths are reported between 9 March and 28 June 2020, which is close to the excess mortality estimated by weekly averages of historical mortality data (8985 deaths). This translates to 837 DPM and an IFR of 1.5% in the general population in Belgium. Both DPM and IFR increase with age and are substantially larger in the nursing home population.

**Conclusion:** Belgium has virtually no discrepancy between COVID-19 reported mortality and excess mortality. Due to this close agreement it is useful to consider the DPM and IFR, which are both age, sex, and nursing home population dependent. Data comparison of COVID-19 mortality between countries should rather be based on excess mortality than reported mortality.

## 1 Introduction

Belgium’s COVID-19 related mortality per million inhabitants has been reported as being the highest worldwide, excluding microstates, over the period 11 April – 26 August 2020. For example, as reported on June 28, 2020 by “Our World in Data” [1], Belgium has 830 COVID-19 Deaths Per Million (DPM), versus 107 for Germany, 379 for the United States (US), 456 for France, 539 for Sweden, 574 for Italy, and 593 for the United Kingdom (UK).

Because of its relative nature, DPM appears to be an objective measure for comparison, but it heavily depends on many factors, including but not limited to the geography and the completeness of COVID-19 mortality reporting. The first half year of 2020, primarily the East Coast of the US was affected by COVID-19, resulting in a relatively low DPM for the entire US. The high death toll at the East Coast is diluted by the largely unaffected West Coast population in the first half year. Indeed, focusing on New York State until the end of June 2020, the DPM is 1599, largely exceeding the Belgian DPM for this period [2]. The completeness of COVID-19 mortality reporting depends on many factors as well, such as directives, availability of data, and the definition of a COVID-19 related death. Belgium is one of the few countries whose COVID-19 mortality notification is broader than the WHO criteria [3] and includes confirmed COVID-19 deaths in nursing homes as well as possible COVID-19 related mortality cases [4, 5]. Because the completeness of reporting is variable between countries and regions, international comparison of COVID-19 reported mortality is biased. Therefore, excess mortality has been recommended as a more reliable and useful metric for comparison [6, 7].

The Case Fatality Rate (CFR), another measure frequently reported regarding COVID-19 mortality, is arguably also not a good basis for international comparison [8, 9]. Besides its dependence on the COVID-19 mortality reporting accuracy, it is also strongly influenced by testing strategy. Additionally, in several studies the delay between case confirmation and death is not accounted for [10] and age dependency is ignored. The handling of suspected COVID-19 cases is ambiguous at best. However, the CFR can be useful as a tool in estimating global Infection Fatality Rates (IFRs) [11], when the IFR is derived as a limit of the CFR by asymptotic models. We do not consider CFR here, but will discuss the IFR.

To understand the subtleties of COVID-19 mortality in Belgium, we examine COVID-19 reported mortality over the period 9 March– 28 June 2020, place it against the background of excess mortality in Belgium, and compare it to the reported mortality of other countries. The study period is chosen to cover the first COVID-19 wave, on which accurate death counts are available following a data cleaning period. This allows us to gauge whether there is evidence for over-, under-or sufficiently accurate reporting of COVID-19 mortality in Belgium. Using data on the number of COVID-19 deaths, COVID-19 hospitalizations and seroprevalence estimates based on data from serial serological surveys [12], the number of deaths per million and infection fatality rates are estimated, overall, and in relation to age and sex, for the general population as a whole, and the nursing home population (NHP), and the non-NHP separately.

## 2 Data and methods

### COVID-19 mortality

The Belgian institute for public health, Sciensano, registers daily COVID-19 mortality figures [13]. These daily data were extracted on 30 September 2020 and then binned to form age category by week mortality tables for each of the sexes and for the period 9 March – 28 June 2020 (Week 11–Week 26); age categories (in years) are 0–9, 10–19, 20–29, 30–39, 40–49, 50–59, 60–69, 70–79, 80–89, 90+. These ten categories are used throughout the analyses, unless otherwise specified. The daily information was binned in Monday to Sunday weeks. Missing data redistribution methods are used to classify all data in an age-week-sex table [14]. Deaths with neither age nor sex observed are redistributed in an ad-hoc fashion over the proper week, so as to match the age-sex distribution observed from historical mortality data. Of the COVID-19 deaths reported in Belgium between 9 March and 28 June 2020, only 1 man is of unknown age, whereas 10 individuals have their age but not their sex reported (all aged 65 years or more), and 15 persons have neither age nor sex available. Although this is a small number of missing observations, redistribution methods were used to impute the missing information. Due to the low amount of missingness, the redistributed data is not influencing the age or sex related results.

In addition, two sub-populations were considered that jointly comprise the NHP deaths: (a) deaths occurring in nursing homes, and (b) nursing home residents that died in hospitals. For the latter category, 129 individuals have missing age and/or sex. Redistribution methods per week are used for deaths with missing information, matching the age-sex distribution observed from the nursing home residents mortality in hospitals in Belgium.

Registered COVID-19 related deaths in Belgium include *confirmed* and *possible* COVID-19 deaths. A case can be confirmed either by a chest CT scan with clinical presentation or a laboratory test. Possible deaths are those who meet the clinical criteria, whether or not there is an epidemiological link to a confirmed case [5, 15].

### General and excess mortality

Weekly mortality per sex and age category, for the years 2009– 2019 (complete) and 2020 (until 28 June 2020) originated from the National Register. Statistics Belgium, the national statistical institute, processed these deaths and integrated them in Demobel, its demographic data warehouse. Open data by district (NUTS 3) can be found in [16]. Using the years 2009–2019 combined, a weekly average profile (termed *baseline*) is obtained, with pointwise corresponding 99% prediction band of a normal distribution. The weekly average profiles were subtracted from the weekly mortality data of 2020 in the period between 9 March–28 June 2020 to estimate the weekly excess mortality for this period, with corresponding 95% Prediction Intervals (PIs).

### Population sizes

The Belgian population sizes (situation 1 January 2020), by age category and sex, were taken from Statistics Belgium (Demobel), based on National Register data [17].

### Estimated number of COVID-19 infected cases

We used a stochastic discrete-time age-structured compartmental model [18] to estimate the number of COVID-19 infected cases. For the purpose of a sensitivity analysis, the number of COVID-19 infected cases was additionally estimated with an individual-based model [19]. Both models are calibrated on national hospitalization data and serial serological survey data [12], while the stochastic model uses additionally Belgian mortality data [13] and the individual-based model employs doubling times [20]. More specifically, the stochastic model was composed of different states including presymptomatic, asymptomatic, and symptomatic compartments and accounts for changes in social contact behavior following the stringent lockdown measures taken and subsequent relaxations thereof. The model predicts (stochastic realizations of) the daily number of new infections per 10 year age groups. The individual-based model, on the other hand, is based on census household data and accounts for presymptomatic, asymptomatic and symptomatic health states in combination with adjusted social contact behavior during the study period. The model output is the daily number of new infections by age, which we aggregate here in the 10 year age groups. The individual-based model was developed to estimate the COVID-19 infections in the general population and to measure the effect of the non-pharmaceutical interventions on the number of infections, it was not developed to estimate the number of infections per age category. Since the individual-based model assumes higher aged individuals to live relatively isolated, in the absence of persons living in collective housings or elderly homes, it is not well-equipped to accommodate outbreaks in nursing homes nor to reliably estimate the number of infections in the 80+ year population. Although the stochastic model does not explicitly accounts for elderly homes, it does allow for substantial transmission in those higher age groups affected by outbreaks and transmission within nursing homes. Hence, it is deemed more reliable with regard to the estimation of the total number of infected cases in the higher age categories.

### Infection fatality rates

Inspired by the work of Nishiura et al. [21], the daily IFR was calculated as the number of deaths on day *t* that were COVID-19 infected, and the total number of infections on day *t*:

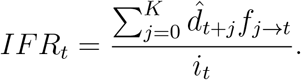

The predicted number of deaths, 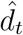, is modeled according to a negative binomial regression, with the mean following a Richards model [22] at time *t* during the study period and beyond (*t* = 0, …, *T*, … *T* + *K*). The delay distribution between infection and death was estimated from the literature and from the individual hospital survey [23], with *f*_*j*_ the probability of a delay of *j* days between infection and death (*j* = 0, …, *K*) and *K* the maximal number of days between infection and death. The time between infection to symptom onset has a lognormal distribution with parameters 1.516 and 0.0164. The time between symptom onset to death was based on a Weibull distribution, which accounted for the interval-censoring nature of the observed delay times and truncation at the end of the study period and is age-specific [23]. Finally, the number of infected cases *i*_*t*_ is the predicted mean number of COVID-19 infections based on the stochastic discrete time age-structured compartmental model [18] or the individual-based model [19].

The posterior distributions of the IFR at each time point were obtained by MCMC sampling of the predicted number of deaths. A summary of the IFR over the time range March 11–June 28 was made by averaging the daily IFR. The 95% confidence interval of the IFR takes account of both the variability of the estimation of the COVID-19 infected cases as the variability of the MCMC sampling. The IFR was estimated per age category, for the general population, the NHP, and the non-NHP.

### Statistical software

The data analysis was performed using SAS Software, GAUSS, and R; visualizations were made using Vega.

### Patient and public involvement

No patient involvement (aggregated mortality data are used).

## 3 Results

### COVID-19 Mortality

Less than 5 COVID-19 deaths occurred in the combined age categories 0–29 years. Due to the low count, these age categories are excluded in the remainder of the analyses. Any measure based on these counts would be highly inaccurate.

Of the 9621 reported COVID-19 deaths, 4535 (5086) are male (female) (Appendix Table 4) and 2591 (27%) are suspected or probable deaths. The majority of the 2591 suspected or probable deaths occurred in a nursing home, i.e., 2310 (89%). In line with international findings [6], the number of deaths strongly increases with age (Figure 1 and Appendix Table 4). The peak of the COVID-19 deaths was reached at week 15, which is the second week of April, starting 6th of April (Figure 1 and Appendix Table 4).

**Figure 1:**
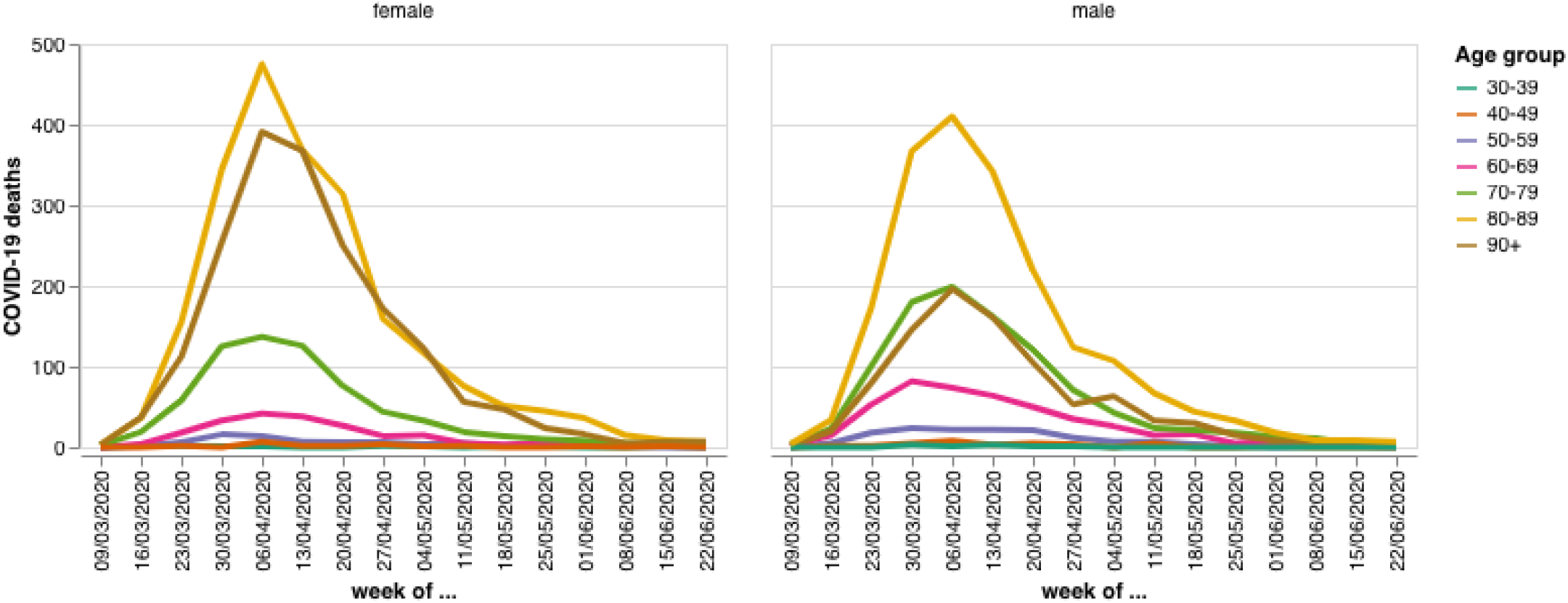
Age group and sex specific COVID-19 mortality in Belgium over the period 9 March–28 June 2020

With 4763 deaths in the nursing homes (15 with missing age and sex, 3 with missing sex) and 1294 nursing home residents who died in hospitals (129 with missing age and/or sex), the majority (63%) of the COVID-19 deaths occurred in the NH population (Appendix Table 5 and 6). It is difficult to compare sexes in absolute terms, because the higher number of deaths in the female 80+ group, for example, is offset by the fact that the number of males in the 80+ category is roughly half the size of the female category (Appendix Table 7). A relative comparison taking account of the size of the population, such as DPM would be more appropriate.

#### Excess deaths

The excess mortality in 2020 is apparent when compared to the first half year of the years 2009–2019 (Figure 2). The mortality until week 10 in 2020 (8 March) was below the baseline (average over years 2009–2019), although coherent with the prediction interval, to rise well over the seasonal variation of the historical mortality data in the subsequent weeks. The mortality peak lies clearly outside the 99% pointwise prediction bands. Although mortality was high in the winter seasons of 2011–2012, 2012–2013, 2014–2015, 2016–2017, and 2017–2018, in the second week of April 2020, twice as many people died than on average over 2009–2019. Looking further into history, April 2020 was the deadliest month of April since World War II, although January 1951 and February 1960 saw similar figures [15, 24].

**Figure 2:**
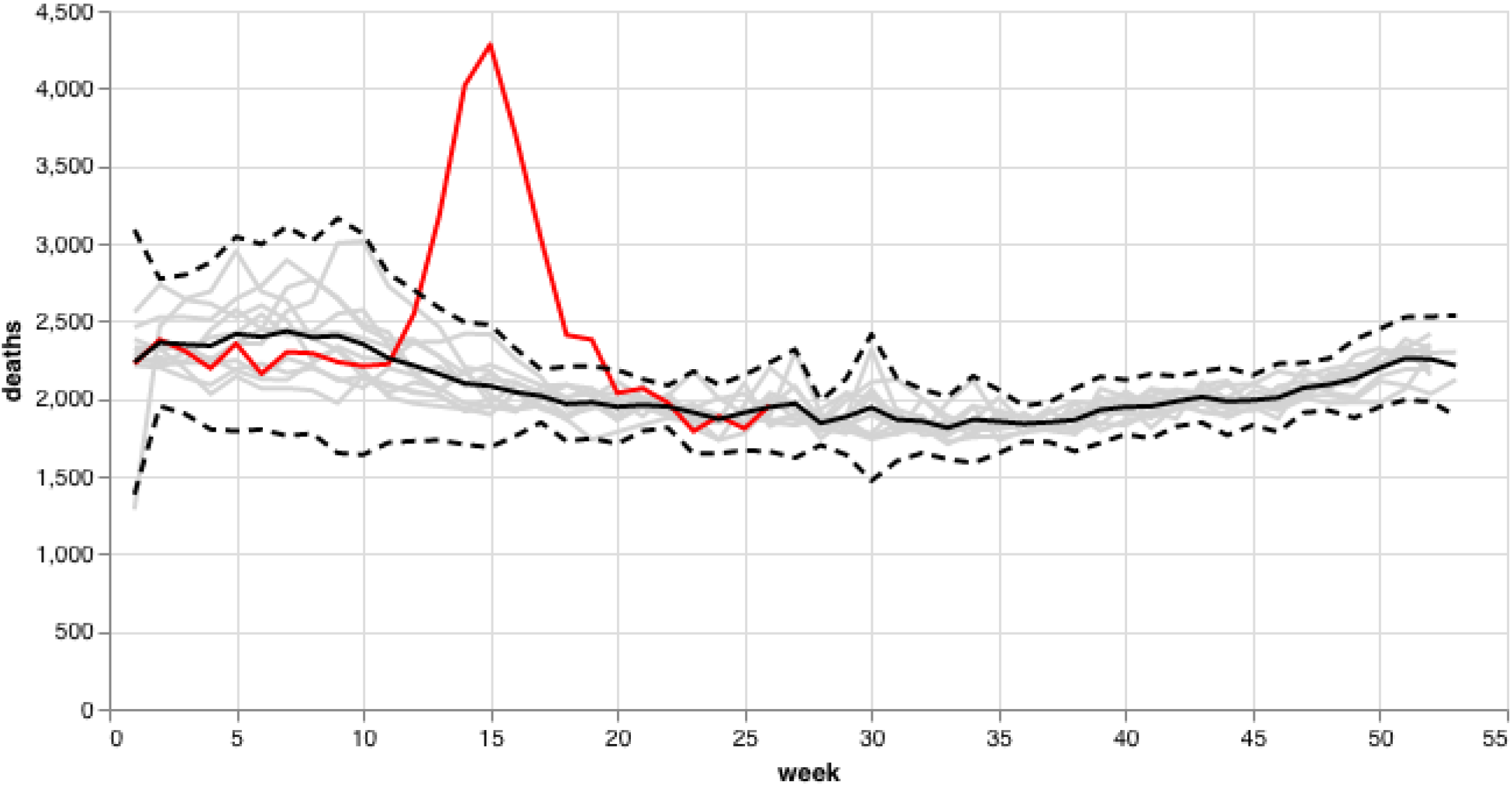
General mortality in Belgium. Grey curves refer to years 2009–2019, with the black curve the average over 2009–2019 and dashed lines the 99% pointwise prediction bands. The red curve refers to year 2020.

The excess mortality in Belgium between week 11 and 26 in 2020 based on the weekly average from 2009–2019 is 8985 (95% PI: 5388-12,582), which is clearly higher than the excess mortality of more recent years. In the winter of 2017–2018, there were 70,215 actual deaths, which is 3093 more than the Belgian Mortality Monitoring (Be-MOMO) model prediction (4.6% excess mortality) [25]. In the winter of 2016–2017, there was an excess of 3284 deaths (4.9% excess mortality) [25].

Notice the near coincidence of the excess and COVID-19 mortality (Figure 3) and that the peak of excess mortality is strongly driven by the older age categories (Appendix Figure 5). The reported COVID-19 deaths as share of excess deaths is 107%, which is slightly different from the 110% reported by Aron et al. [6], because a thorough revision of the reported mortality cases occurred after the publication of the paper and because we consider additional weeks and additional historical data.

**Figure 3:**
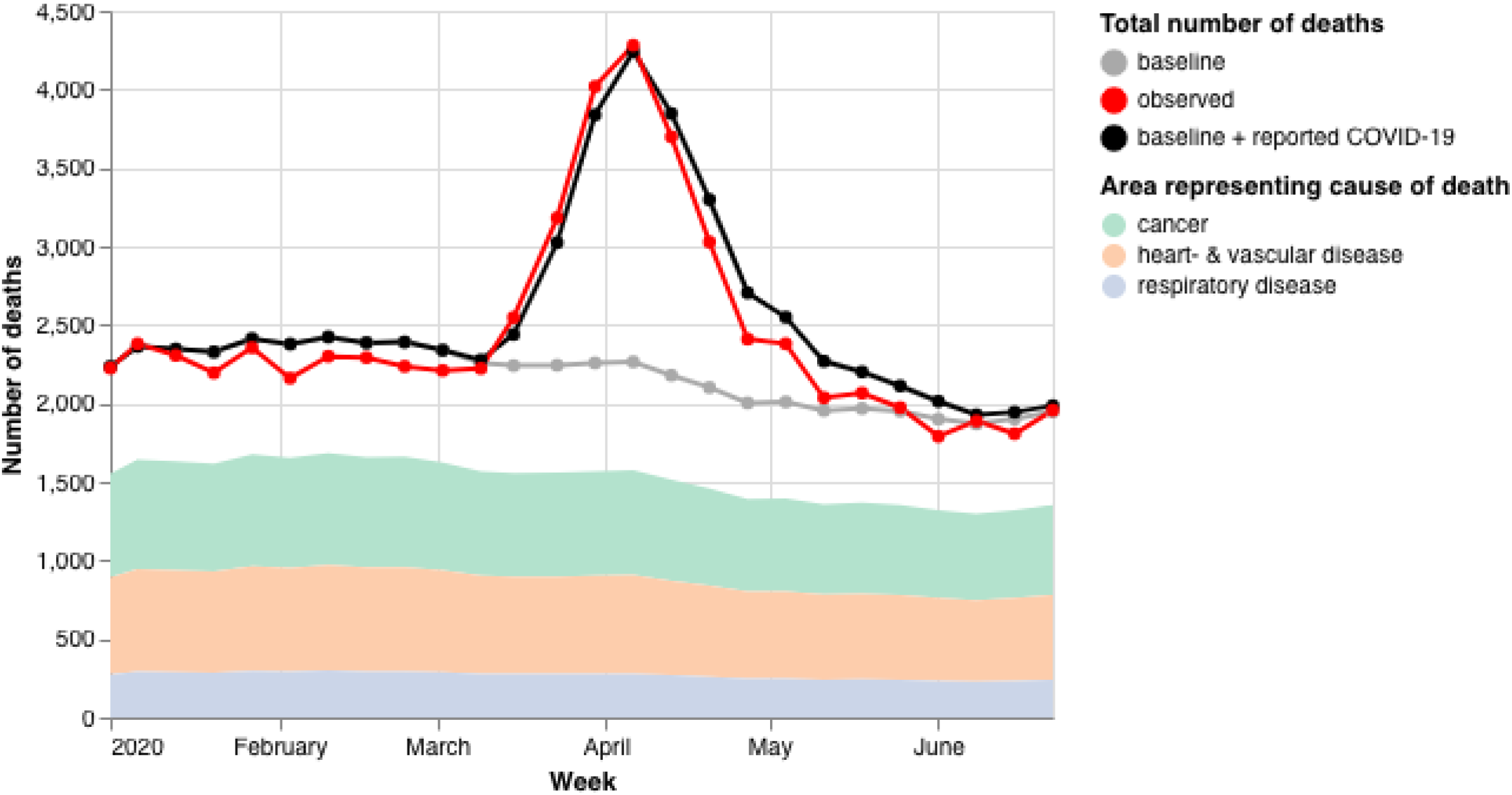
Observed general mortality in 2020 (red curve) versus the sum (black curve) of reported COVID-19 mortality and the average of 2009–2019 mortality (grey curve).

**Figure 4:**
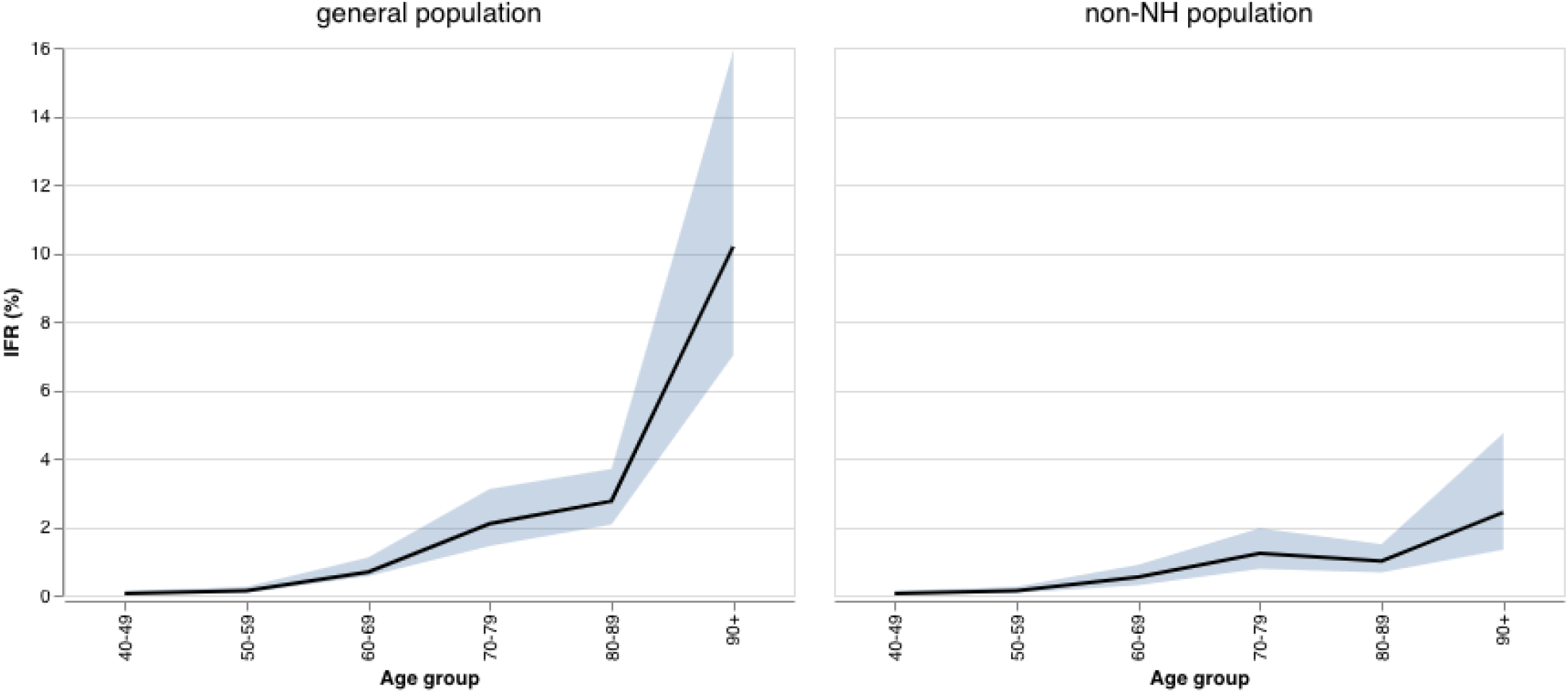
Infection fatality rate (%) in the general population (left) and the non-NH population (right).

**Figure 5:**
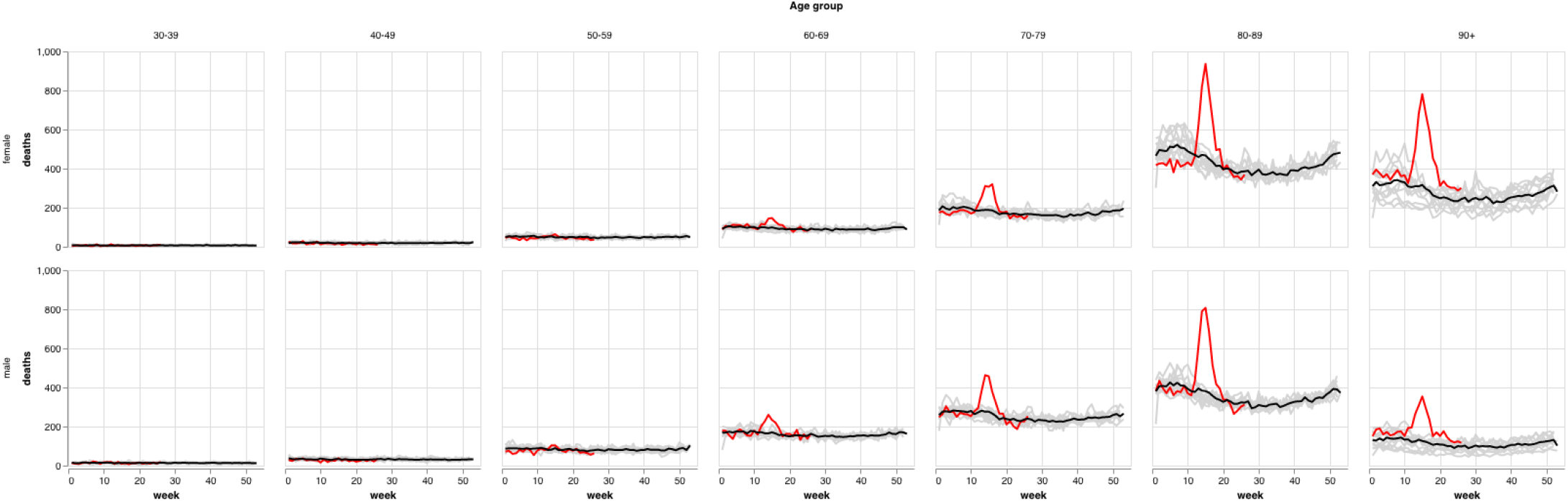
General mortality by age group and gender in Belgium. Grey curves refer to years 2009– 2019, with the black curve the average over 2009– 2019. The red curve refers to year 2020.

#### Covid-19 Number of Deaths per Million Inhabitants

Now that the close agreement between COVID-19 and excess mortality has been established, it is useful to consider the number of deaths per million inhabitants. Belgium counts over the study period 837 DPM (Table 1), which differs slightly from the 830 DPM reported by “Our World in Data” [1] (Table 2). Again this difference can be explained by the revision of the reported mortality.

**Table 1:** Number of COVID-19 deaths per million inhabitants in Belgium per age category and sex for the non-NH, NH and general population.

| Age |  | 25–49 | 50–59 | 60–69 | 70–79 | 80–89 | 90+ | All Ages combined | Combined over 60+ strata |
| --- | --- | --- | --- | --- | --- | --- | --- | --- | --- |
| Female | non-NH | 19 | 93 | 200 | 654 | 1834 | 4349 | 318 | 876 |
|  | NH |  |  | 25379 | 34408 | 41673 | 53604 | 44486 | 44486 |
|  | General population | 19 | 92 | 311 | 1372 | 6743 | 22110 | 872 | 3183 |
| Male | non-NH | 25 | 182 | 538 | 1431 | 4081 | 10787 | 563 | 1720 |
|  | NH |  |  | 29463 | 61234 | 91117 | 98069 | 78751 | 78751 |
|  | General population | 25 | 182 | 687 | 2343 | 9305 | 28201 | 801 | 3409 |
| Both sexes | non-NH | 22 | 138 | 365 | 1014 | 2753 | 6449 | 438 | 1266 |
|  | NH |  |  | 27391 | 44633 | 53495 | 61464 | 53267 | 53267 |
|  | General population | 22 | 138 | 495 | 1821 | 7748 | 23808 | 837 | 3286 |

**Table 2:** Ranking of countries by deaths per million and adjusted deaths per million on 28 June 2020

| Country | Deaths Per Million | Country | Adjusted Deaths Per Million |
| --- | --- | --- | --- |
| **Belgium** | 830 | New York City | 2222 |
| Spain | 606 | New York State | 1599 |
| United Kingdom | 593 | Spain | 1010 |
| Italy | 574 | Italy | 857 |
| Sweden | 539 | **Belgium** | 755 |
| France | 456 | United Kingdom | 742 |
| United States | 379 | Netherlands | 574 |
| Netherlands | 356 | France | 470 |
| Germany | 107 |  |  |

Source: Our World in Data [1]

Similar to the reported COVID-19 mortality data, a strong age and important sex effect is observed in the DPM (Table 1). The DPM increases exponentially with increasing age, while in all age categories the male DPM is higher than the female. But what is most striking is the extent of the impact in the NHP. For the non-NHP, the overall figure goes down to 438 deaths per million, which internationally might not stand out, but then reliable figures are needed of other countries’ NHP/non-NHP deaths as well. For a nursing home population of around 1% of the total population, this effect is striking.

It is clear from Table 1 that the overall DPM is not very informative, but rather an age, sex, and population-specific breakout is necessary.

When comparing the deaths per million between countries, the different degree of reporting of COVID-19 mortality by the countries should be acknowledged. For example, considering the ratio of the reported COVID-19 mortality and the excess mortality reported in [6], Belgium’s adjusted DPM of 755 is comparable to the UK’s.

### Infection Fatality Rates

The number of COVID-19 infections in Belgium estimated by the stochastic model and the individual-based model are similar for the lower age categories (Appendix Figure 6), while in the upper categories, as expected, they disagree. Using the number of daily estimated COVID-19 infections and daily COVID-19 mortality with a delay distribution, the daily IFR was estimated.

**Figure 6:**
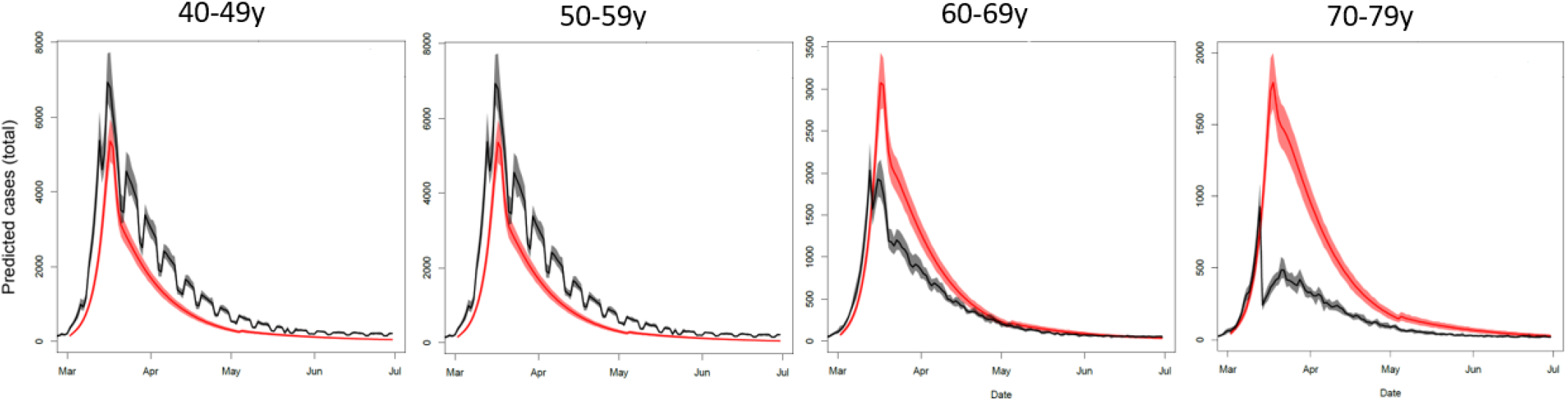
Estimated number of COVID-19 infections by age group in Belgium. Red curves refer to the estimation by the stochastic compartmental model and black curves to the estimation by the individual-based model.

Based on the stochastic model, the IFR across all ages is estimated at 1.5% in the general population (Table 3). The IFR shows an age dependent exponential increase, with nearly 0% under 40 years, to 10% above 89 years in the general population (Figure 4). Compared to the meta-analysis of Levin et al. [26], 0.4% at age 55, 1.3% at age 65, 4.2% at age 75, 14% at age 85, and *>*25% for ages 90 and above, we find lower IFRs in the 70+ population.

**Table 3:** Infection fatality rate in Belgium with 95% confidence interval per age category for the non-NH, NH and the general population with the stochastic model.

|  | Age |  |  |  |  |  | All Ages combined |
| --- | --- | --- | --- | --- | --- | --- | --- |
|  | 40–49 | 50–59 | 60–69 | 70–79 | 80–89 | 90+ |  |
| non-NH | 0.05<br>(0.01–0.13) | 0.14<br>(0.07–0.26) | 0.53<br>(0.30–0.90) | 1.23<br>(0.78–1.96) | 1.00<br>(0.67–1.50) | 2.42<br>(1.34–4.73) | 0.58<br>(0.42–0.81) |
| NH |  |  | 31.42<br>(15.43–63.13) | 45.91<br>(29.89–72.37) | 18.46<br>(13.55–25.36) | 26.27<br>(17.90–41.26) | 20.98<br>(15.83–28.58) |
| General population | 0.05<br>(0.01–0.13) | 0.14<br>(0.07–0.26) | 0.68<br>(0.56–1.11) | 2.09<br>(1.44–3.11) | 2.75<br>(2.07–3.69) | 10.18<br>(7.01–15.90) | 1.47<br>(1.14–1.94) |

The striking difference between the COVID-19 mortality in the non-NHP and the NHP seen in the DPM, also shows in the IFR, with 0.6% and 21% respectively. These estimates of the IFR are very similar to ones reported for France by O’Driscoll et al. [27], non-NHP 0.7%, NHP 22.3% and 1.1% in the general population. Depending on the age, the IFR in the NHP is 10 to 60 fold higher than in the non-NHP. Interestingly, when comparing the ratio of the IFR or DPM in the NHP versus the non-NHP within an age group, the ratio decreases with increasing age. This may suggest that compared to the older NH residents, the younger NH residents are more frail compared to the non-NH individuals of the same age group.

The IFRs from the individual-based model coincide well with the IFRs from the stochastic model when looking across all ages in the general population and the non-NHP (Appendix Table 8). As expected for the very high age groups and in the NHP, the individual-based model overestimates the IFR.

## 4 Discussion

When it comes to COVID-19 mortality, Belgium is often cited as one of the worst countries worldwide. Therefore, we studied in detail the COVID-19 mortality, excess mortality and its relation to DPM and IFR in Belgium in the first half year of 2020 in age and NH dependent subgroups and placed them in perspective of internationally reported COVID-19 mortality.

The COVID-19 mortality in Belgium suggests the seriousness of the epidemic. Belgium’s April 2020 mortality was the highest among all months of April since World War II. In the second week of April 2020, COVID-19 mortality was twice as high as long-term average-mortality for that week, exceeding by far the influenza-related increases of mortality of the recent 10 years. The epidemic affected severely NH residents in Belgium, which is only partly explained by the association between age and COVID-19 mortality.

Belgium decided early in the pandemic to report not only COVID-19 laboratory test or chest CT scan confirmed deaths, but also possible COVID-19 cases [5]. Because the COVID-19 mortality monitoring varies between countries, due to several reasons, international comparisons may be seriously biased [7]. The close agreement between COVID-19 mortality reported deaths in Belgium and the excess mortality, pleads in favor of the reporting strategy in Belgium [15]. This coincidence is however not a proof for the fact that all excess deaths are COVID-19 related, although it has been reported internationally that around 90% of suspected deaths are proper COVID-19 deaths [11]. While it may be possible, for example, that some excess deaths are related to other factors, such as lockdown-induced stress, the plausible assumption is made that this effect on mortality is minor [7]. Further examination is warranted as soon as the cause-specific mortality database becomes available, typically after a three-year interval.

The difference between excess mortality and COVID-19 mortality reported deaths in Belgium, may be due to the inaccuracy of predicting the 2020 mortality with the weekly averaging method [7]. Accurate reporting of COVID-19 mortality should also be higher than registered excess mortality. Indeed, in frail and aged individuals, COVID-19 may be a competing risk for other fatal conditions, such as heart failure or cerebrovascular accident. Since some of these individuals may have died in the first half of 2020 anyway of these other fatal conditions, they are counted, rightfully so, as a COVID-19 related death, but do not contribute to the excess mortality.

It is difficult to compare COVID-19 mortality to countries that have a less extensive reporting strategy, in particular when the gap between excess deaths and COVID-19 mortality is large, such as in the Netherlands, Italy, or Austria [2]. Currently, EuroMOMO allows for comparisons by means of the Z-score, a useful metric that indicates how unusual mortality is over a given period, relative to average mortality in that same period. However, it has been pointed out that Z-scores are strictly speaking not comparable between countries [6]. For example, countries with less extreme variations in the reference period will have a smaller variance and for the same deviation in the epidemic period a larger Z, compared to a country with more variation in the reference period. It may thus be useful to supplement it with other metrics. Arguably, excess mortality is a better basis for international comparison [6, 7].

Obviously, the mortality related measures, DPM and IFR, are dependent on the completeness of the mortality reporting. If one wants to compare COVID-19 mortality between countries, it would be better to take account of the possible under-reporting of mortality cases and adjust the DPM for example by using estimates of the amount of under-reporting. Doing so, Belgium’s COVID-19 mortality would still be high, but no longer an extreme.

Despite the completeness of mortality reporting, the DPM and IFR depend on other important factors. Although DPM and IFR clearly increase with age, adjusting for age distribution of a country is of less importance when comparing European and Western countries. Despite differences in the age distribution between European countries, Canada and the US [28], most fluctuate around a proportion of 20% of the population above 65 years. The age distribution would be more important when comparing Western versus African or Asian countries. A higher proportion of older people may have various demographic reasons, such as low fertility during a few years, migration and increasing general level of well-being. The role of high-quality health care facilities on the demographic age distribution is more debated, but with increasing aged population, underlying comorbidities such as high blood pressure and diabetes are more prevalent, which are known to be risk factors for COVID-19 mortality. This is possibly explaining partly the increased mortality with age.

Directly related to this, but worth separate mention, is that the epidemic has been very severe in the NHP in Belgium, which shows in both the DPM and IFR, suggesting a nuanced explanation. The age decrease DPM and IFR ratio between the NHP and non-NHP COVID-19 mortality for a given age, suggests a larger difference in frailty and prevalence of underlying comorbidities in the 60–79 years age group than the 80+ age group. It is indeed plausible to assume that if someone requires the nursing and caring attention in a NH at age 60, that he or she have some limiting comorbidities or increased frailty. In the NHP, besides the health status, also, the effect of vectors, such as caregivers, should not be underestimated and protection and preventive measures taken in view of possible future outbreaks. In summary, the very large DPM in the NHP versus the non-NHP, when compared within a given age, arguably results from a larger number of infections, in combination with an increased IFR. All in all, the outbreak in Belgian nursing homes was extremely serious, in line with international findings [29, 30].

Between European and Western countries, many other factors influence the DPM and IFR, such as

1. International connectivity and internal patterns. Besides an international well-connected airport and multiple entry points, in Belgium an important fraction of the population commutes for work or study related reasons. Additionally, events such as carnival festivities or individuals returning from ski holidays abroad may have influenced the first COVID-19 outbreak. Spring break in Belgium took place in week 9 for the entire country, whereas in some countries it is spread over two (the Netherlands) or four (Sweden) weeks. Belgium had several clusters simultaneously whereas, for example, in the Netherlands the virus was introduced in the south while the north was relatively spared. The presence of communities with Italian heritage that travelled back from spring breaks is an additional contributing factor.
2. The population density in a country, which depends among other on the size and geographical dispersion of a country. Large countries with loosely connected regions and/or low population density might see a much slower increase of seroprevalence but with larger regional differences, than a compact and well-connected country. For example, Sweden’s population density is 15 times smaller than Belgium’s. Also, the presence of large shopping centers and housing situation impact the spread of an infection. Areas with large apartment buildings and households sharing a house will be more prone to a high increase in seroprevalence.
3. The timing of the epidemic. The mortality should be compared relative to a well-defined baseline (e.g., 50 days since the first day at which the DPM exceeded 1.0) rather than calendar time. This would produce, for example: Italy, 24 April, 423; France, 7 May, 443; UK, 7 May, 443; Belgium, 8 May, 726; Sweden, 10 May, 319; US, 11 May, 240; and Germany, 12 May, 90.
4. The varying measures taken by national and regional authorities to fight the epidemic.

Although the IFR is a useful and interesting measure to compare COVID-19 mortality, the number of COVID-19 infections is an additional source for bias and uncertainty. This suggests the use of statistical sensitivity analysis, by applying different methods to estimate the number of infected cases, along with the reporting of interval estimates. The methods used to estimate the number of infected cases in our analysis could potentially be further improved by using seroprevalence data specifically for the NH population. This data is largely unavailable at the moment. The compartmental and individual-based models used to estimate the number of COVID-19 infections now assume a similar seroprevalence in the NH population as the non-NH population.

For the general population, the steep age-related gradient in mortality, expressed in IFR or DPM, contributes useful information to policymakers for differential non-pharmaceutical interventions. However, a more detailed study and further international comparison of COVID-19 mortality is urgent, as well as the implementation of targeted non-pharmaceutical interventions, while awaiting promising pharmaceutical development.

## Data Availability

Data can be obtained from the authors upon reasonable request.

## Contributorship statement

Conceptualization of project: GM, CF, NH, PVD, FR, NBS, HVO, BD, PD; initial draft: GM; editing: all authors; input regarding various data types: (a) general population and overall mortality: PL, NBS; (b) seroprevalences: SH, SA, HT, PVD, NH; (c) COVID-19 mortality: FR, JVDH, NBS, BD, HVO; data analysis: (a) overall: GM, JV; (b) seroprevalence: SH; (c) incidence of infections: SA, LW; (d) infection fatality rates: CF. The investigators were independent from the funders; all authors had full access to the data and can take responsibility for the integrity of the data and the accuracy of the data analysis; the lead author affirms that the manuscript is an honest, accurate, and transparent account of the study being reported; that no important aspects of the study have been omitted. This works reflects the views of the authors and not necessarily the official position of the institutions they belong to.

## Data sharing statement

Data and software code used for the tabular and graphical displays in this study are publicly available from https://www.uhasselt.be/DSI.

## Acknowledgements

We are grateful for the ability to use open data on COVID-19 mortality and cause-specific mortality (Sciensano, Belgium), general mortality and population figures (Statistics Belgium, Demobel; National Register). The data providers hold no responsibility for the analyses reported in this manuscript. We thank Sciensano colleagues Sophie Quoilin, Katrien Tersago, Dominique Van Beckhoven, Nina Van Goethem, and others, for suggesting relevant data sources from among their publicly available data, for useful comments and critical reflections on the analysis strategy, and for comments on earlier drafts of the manuscript. Particular thanks go out to Sciensano colleagues Sara Dequeker and Eline Vandael from the Nursing Homes Surveillance Team. First author GM thanks Barbara Debusschere for suggesting an earlier version of this project. Finally, our gratitude goes to the reviewers, their thoughtful suggestions were invaluable to improve an earlier version of the manuscript.

### A Appendices

**Table 4:** COVID-19 mortality in the general population in Belgium per age category, sex and week

| Age | Week |  |  |  |  |  |  |  |  |  |  |  |  |  |  |  | Total |
| --- | --- | --- | --- | --- | --- | --- | --- | --- | --- | --- | --- | --- | --- | --- | --- | --- | --- |
|  | 11 | 12 | 13 | 14 | 15 | 16 | 17 | 18 | 19 | 20 | 21 | 22 | 23 | 24 | 25 | 26 |  |
| Male |  |  |  |  |  |  |  |  |  |  |  |  |  |  |  |  |  |
| 30–39 | 0 | 0 | 0 | 3 | 1 | 3 | 1 | 1 | 0 | 0 | 0 | 0 | 0 | 0 | 0 | 0 | 9 |
| 40–49 | 0 | 2 | 2 | 5 | 8 | 3 | 5 | 4 | 0 | 5 | 0 | 0 | 1 | 0 | 0 | 0 | 35 |
| 50–59 | 0 | 5 | 18 | 24 | 22 | 22 | 21 | 12 | 7 | 7 | 4 | 2 | 0 | 1 | 1 | 0 | 146 |
| 60–69 | 2 | 15 | 53 | 82 | 74 | 64 | 50 | 35 | 26 | 15 | 16 | 5 | 8 | 4 | 2 | 2 | 453 |
| 70–79 | 4 | 23 | 101 | 180 | 199 | 162 | 121 | 71 | 43 | 24 | 21 | 18 | 13 | 11 | 4 | 6 | 1001 |
| 80–89 | 4 | 34 | 174 | 367 | 410 | 342 | 219 | 124 | 107 | 67 | 44 | 33 | 18 | 9 | 9 | 7 | 1968 |
| 90+ | 1 | 20 | 80 | 146 | 196 | 161 | 105 | 53 | 63 | 33 | 30 | 15 | 8 | 5 | 3 | 4 | 923 |
| Total | 11 | 99 | 428 | 807 | 910 | 757 | 522 | 300 | 246 | 151 | 115 | 73 | 48 | 30 | 19 | 19 | 4535 |
| Female |  |  |  |  |  |  |  |  |  |  |  |  |  |  |  |  |  |
| 30–39 | 0 | 2 | 2 | 1 | 1 | 0 | 0 | 2 | 1 | 0 | 1 | 1 | 0 | 0 | 1 | 0 | 12 |
| 40–49 | 0 | 0 | 2 | 0 | 7 | 2 | 2 | 4 | 1 | 2 | 0 | 0 | 1 | 0 | 1 | 0 | 22 |
| 50–59 | 0 | 1 | 6 | 16 | 14 | 7 | 6 | 6 | 4 | 5 | 2 | 3 | 2 | 1 | 0 | 0 | 73 |
| 60–69 | 0 | 4 | 18 | 33 | 42 | 38 | 27 | 14 | 15 | 5 | 3 | 5 | 2 | 2 | 4 | 2 | 214 |
| 70–79 | 2 | 19 | 58 | 125 | 137 | 126 | 76 | 44 | 33 | 19 | 14 | 10 | 8 | 5 | 2 | 4 | 682 |
| 80–89 | 3 | 36 | 155 | 344 | 475 | 369 | 314 | 159 | 117 | 76 | 51 | 45 | 36 | 15 | 9 | 7 | 2211 |
| 90+ | 3 | 37 | 112 | 254 | 391 | 368 | 250 | 172 | 123 | 56 | 47 | 24 | 16 | 5 | 7 | 7 | 1872 |
| Total | 8 | 99 | 353 | 773 | 1067 | 910 | 675 | 401 | 294 | 163 | 118 | 88 | 65 | 28 | 24 | 20 | 5086 |
| Male+Female |  |  |  |  |  |  |  |  |  |  |  |  |  |  |  |  |  |
| 30–39 | 0 | 2 | 2 | 4 | 2 | 3 | 1 | 3 | 1 | 0 | 1 | 1 | 0 | 0 | 1 | 0 | 21 |
| 40–49 | 0 | 2 | 4 | 5 | 15 | 5 | 7 | 8 | 1 | 7 | 0 | 0 | 2 | 0 | 1 | 0 | 57 |
| 50–59 | 0 | 6 | 24 | 40 | 36 | 29 | 27 | 18 | 11 | 12 | 6 | 5 | 2 | 2 | 1 | 0 | 219 |
| 60–69 | 2 | 19 | 71 | 115 | 116 | 102 | 77 | 49 | 41 | 20 | 19 | 10 | 10 | 6 | 6 | 4 | 667 |
| 70–79 | 6 | 42 | 159 | 305 | 336 | 288 | 197 | 115 | 76 | 43 | 35 | 28 | 21 | 16 | 6 | 10 | 1683 |
| 80–89 | 7 | 70 | 329 | 711 | 885 | 711 | 533 | 283 | 224 | 143 | 95 | 78 | 54 | 24 | 18 | 14 | 4179 |
| 90+ | 4 | 57 | 192 | 400 | 587 | 529 | 355 | 225 | 186 | 89 | 77 | 39 | 24 | 10 | 10 | 11 | 2795 |
| Total | 19 | 198 | 781 | 1580 | 1977 | 1667 | 1197 | 701 | 540 | 314 | 233 | 161 | 113 | 58 | 43 | 39 | 9621 |

**Table 5:** COVID-19 mortality in the nursing home population in Belgium per age category, sex and week

| Age | Week |  |  |  |  |  |  |  |  |  |  |  |  |  |  |  | Total |
| --- | --- | --- | --- | --- | --- | --- | --- | --- | --- | --- | --- | --- | --- | --- | --- | --- | --- |
|  | 11 | 12 | 13 | 14 | 15 | 16 | 17 | 18 | 19 | 20 | 21 | 22 | 23 | 24 | 25 | 26 |  |
| Male |  |  |  |  |  |  |  |  |  |  |  |  |  |  |  |  |  |
| 60–69 | 0 | 0 | 10 | 16 | 30 | 17 | 10 | 2 | 5 | 2 | 3 | 1 | 3 | 1 | 0 | 0 | 101 |
| 70–79 | 0 | 5 | 26 | 73 | 100 | 71 | 48 | 28 | 18 | 11 | 6 | 2 | 5 | 4 | 1 | 0 | 399 |
| 80–89 | 3 | 10 | 55 | 191 | 295 | 221 | 138 | 74 | 68 | 36 | 30 | 18 | 7 | 4 | 4 | 2 | 1157 |
| 90+ | 0 | 10 | 36 | 98 | 138 | 126 | 77 | 44 | 53 | 23 | 15 | 10 | 4 | 1 | 2 | 3 | 640 |
| Total | 3 | 26 | 128 | 379 | 563 | 434 | 273 | 148 | 144 | 72 | 54 | 31 | 19 | 10 | 7 | 5 | 2297 |
| Female |  |  |  |  |  |  |  |  |  |  |  |  |  |  |  |  |  |
| 60–69 | 0 | 1 | 2 | 12 | 18 | 20 | 8 | 3 | 6 | 2 | 1 | 0 | 2 | 0 | 2 | 0 | 77 |
| 70–79 | 0 | 3 | 16 | 56 | 90 | 79 | 46 | 23 | 22 | 7 | 9 | 3 | 4 | 2 | 1 | 2 | 364 |
| 80–89 | 1 | 24 | 76 | 247 | 393 | 303 | 255 | 126 | 89 | 59 | 41 | 28 | 23 | 8 | 7 | 3 | 1684 |
| 90+ | 0 | 28 | 75 | 214 | 357 | 319 | 227 | 164 | 104 | 57 | 38 | 22 | 15 | 4 | 7 | 6 | 1636 |
| Total | 1 | 56 | 169 | 530 | 858 | 722 | 536 | 316 | 221 | 125 | 89 | 53 | 44 | 14 | 17 | 11 | 3761 |
| Male+Female |  |  |  |  |  |  |  |  |  |  |  |  |  |  |  |  |  |
| 60–69 | 0 | 1 | 12 | 29 | 48 | 37 | 18 | 5 | 11 | 4 | 4 | 1 | 5 | 1 | 2 | 0 | 177 |
| 70–79 | 0 | 9 | 42 | 130 | 190 | 150 | 94 | 51 | 40 | 18 | 15 | 5 | 9 | 6 | 2 | 2 | 763 |
| 80–89 | 4 | 34 | 131 | 438 | 688 | 524 | 394 | 200 | 157 | 95 | 71 | 46 | 30 | 12 | 11 | 5 | 2841 |
| 90+ | 0 | 38 | 111 | 312 | 495 | 445 | 304 | 208 | 157 | 80 | 53 | 32 | 19 | 5 | 9 | 9 | 2277 |
| Total | 4 | 82 | 296 | 909 | 1420 | 1156 | 809 | 464 | 365 | 197 | 143 | 84 | 63 | 24 | 24 | 16 | 6057 |

**Table 6:** COVID-19 mortality in the non-nursing home population in Belgium per age category, sex and week

| Age | Week |  |  |  |  |  |  |  |  |  |  |  |  |  |  |  | Total |
| --- | --- | --- | --- | --- | --- | --- | --- | --- | --- | --- | --- | --- | --- | --- | --- | --- | --- |
|  | 11 | 12 | 13 | 14 | 15 | 16 | 17 | 18 | 19 | 20 | 21 | 22 | 23 | 24 | 25 | 26 |  |
| Male |  |  |  |  |  |  |  |  |  |  |  |  |  |  |  |  |  |
| 30–39 | 0 | 0 | 0 | 3 | 1 | 3 | 1 | 1 | 0 | 0 | 0 | 0 | 0 | 0 | 0 | 0 | 9 |
| 40–49 | 0 | 2 | 2 | 5 | 8 | 3 | 5 | 4 | 0 | 5 | 0 | 0 | 1 | 0 | 0 | 0 | 35 |
| 50–59 | 0 | 5 | 18 | 24 | 22 | 22 | 21 | 12 | 7 | 7 | 4 | 2 | 0 | 1 | 1 | 0 | 146 |
| 60–69 | 2 | 15 | 43 | 66 | 44 | 47 | 41 | 33 | 21 | 13 | 13 | 4 | 5 | 3 | 2 | 2 | 352 |
| 70–79 | 4 | 18 | 75 | 107 | 99 | 91 | 73 | 43 | 25 | 13 | 15 | 16 | 8 | 7 | 3 | 6 | 602 |
| 80–89 | 1 | 24 | 119 | 176 | 115 | 121 | 81 | 50 | 39 | 31 | 14 | 15 | 11 | 5 | 5 | 5 | 811 |
| 90+ | 1 | 10 | 44 | 48 | 58 | 35 | 28 | 9 | 10 | 10 | 15 | 5 | 4 | 4 | 1 | 1 | 283 |
| Total | 8 | 73 | 300 | 428 | 347 | 323 | 249 | 152 | 102 | 79 | 61 | 42 | 29 | 20 | 12 | 14 | 2238 |
| Female |  |  |  |  |  |  |  |  |  |  |  |  |  |  |  |  |  |
| 30–39 | 0 | 2 | 2 | 1 | 1 | 0 | 0 | 2 | 1 | 0 | 1 | 1 | 0 | 0 | 1 | 0 | 12 |
| 40–49 | 0 | 0 | 2 | 0 | 7 | 2 | 2 | 4 | 1 | 2 | 0 | 0 | 1 | 0 | 1 | 0 | 22 |
| 50–59 | 0 | 1 | 6 | 16 | 14 | 7 | 6 | 6 | 4 | 5 | 2 | 3 | 2 | 1 | 0 | 0 | 73 |
| 60–69 | 0 | 3 | 16 | 21 | 24 | 18 | 19 | 11 | 9 | 3 | 2 | 5 | 0 | 2 | 2 | 2 | 137 |
| 70–79 | 2 | 16 | 42 | 69 | 47 | 47 | 30 | 21 | 11 | 12 | 5 | 7 | 4 | 3 | 1 | 2 | 318 |
| 80–89 | 2 | 12 | 79 | 97 | 82 | 66 | 59 | 33 | 28 | 17 | 10 | 17 | 13 | 7 | 2 | 4 | 527 |
| 90+ | 3 | 9 | 37 | 40 | 34 | 49 | 23 | 8 | 19 | -1 | 9 | 2 | 1 | 1 | 0 | 1 | 236 |
| Total | 7 | 43 | 185 | 243 | 209 | 188 | 139 | 85 | 73 | 38 | 29 | 35 | 21 | 14 | 7 | 9 | 1325 |
| Male+Female |  |  |  |  |  |  |  |  |  |  |  |  |  |  |  |  |  |
| 30–39 | 0 | 2 | 2 | 4 | 2 | 3 | 1 | 3 | 1 | 0 | 1 | 1 | 0 | 0 | 1 | 0 | 21 |
| 40–49 | 0 | 2 | 4 | 5 | 15 | 5 | 7 | 8 | 1 | 7 | 0 | 0 | 2 | 0 | 1 | 0 | 57 |
| 50–59 | 0 | 6 | 24 | 40 | 36 | 29 | 27 | 18 | 11 | 12 | 6 | 5 | 2 | 2 | 1 | 0 | 219 |
| 60–69 | 2 | 18 | 59 | 86 | 68 | 65 | 60 | 44 | 30 | 16 | 15 | 9 | 5 | 5 | 4 | 4 | 490 |
| 70–79 | 6 | 33 | 117 | 175 | 146 | 138 | 103 | 64 | 36 | 25 | 20 | 23 | 12 | 10 | 4 | 8 | 920 |
| 80–89 | 3 | 36 | 198 | 273 | 197 | 187 | 139 | 83 | 67 | 48 | 24 | 32 | 24 | 12 | 7 | 9 | 1339 |
| 90+ | 4 | 19 | 81 | 88 | 92 | 84 | 51 | 17 | 29 | 9 | 24 | 7 | 5 | 5 | 1 | 2 | 518 |
| Total | 15 | 116 | 485 | 671 | 557 | 511 | 388 | 237 | 175 | 117 | 90 | 77 | 50 | 34 | 19 | 23 | 3564 |

**Table 7:**
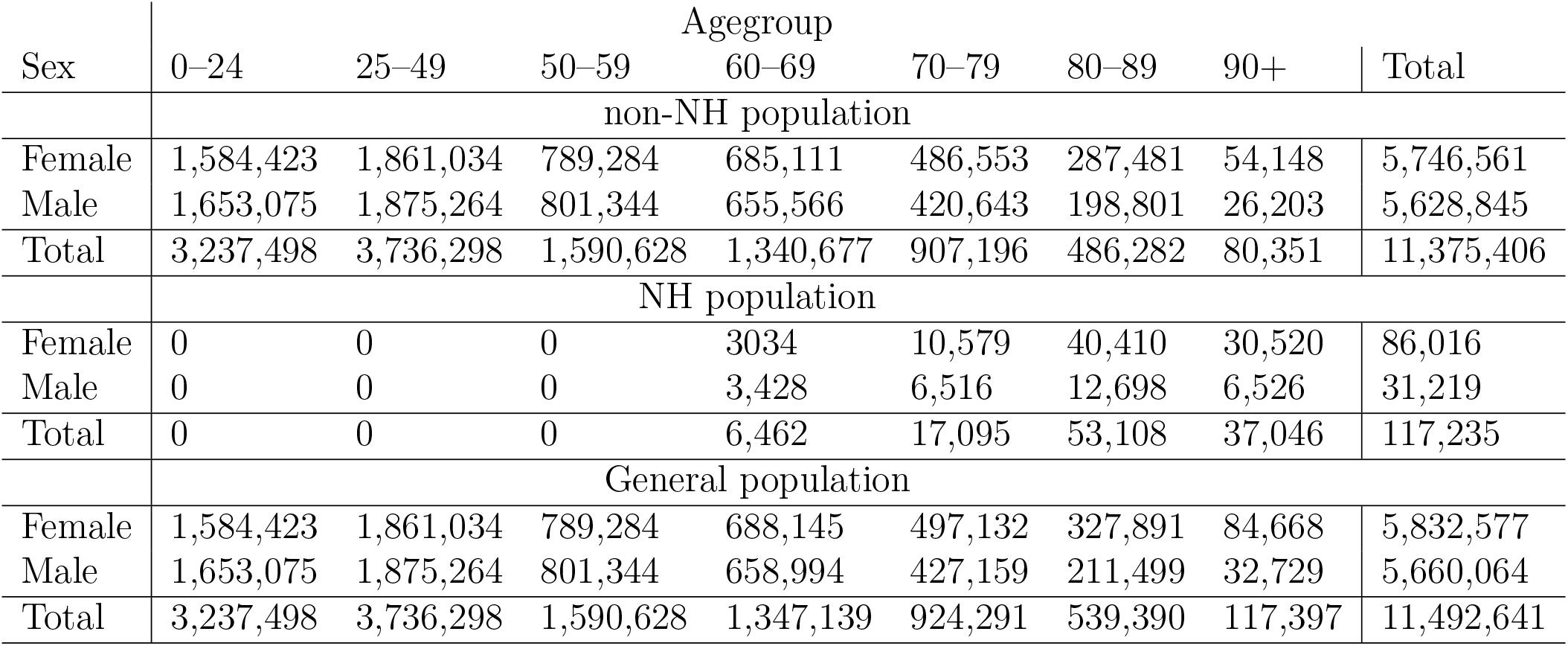
Population of Belgium per age category and sex

**Table 8:** Infection fatality rate in Belgium with 95% confidence interval per age category for the non-NH and the general population with the individual-based model.

|  | Age |  |  |  | All Ages combined |
| --- | --- | --- | --- | --- | --- |
|  | 40–49 | 50–59 | 60–69 | 70–79 |  |
| non-NH | 0.03<br>(0.01–0.06) | 0.12<br>(0.06–0.20) | 0.61<br>(0.38–0.95) | 2.16<br>(1.42–3.30) | 0.65<br>(0.50–0.84) |
| General population | 0.03<br>(0.01–0.06) | 0.12<br>(0.06–0.20) | 0.79<br>(0.52–1.18) | 3.72<br>(2.64–5.35) | 1.72<br>(1.39–2.12) |

